# Dysmorphological and neuropsychological phenotypes of prenatally alcohol-exposed six-year-old children

**DOI:** 10.1101/2025.10.28.25338948

**Authors:** M Jolma, E Wallén, E Saure, K Rämö, H Kahila, N Kaminen-Ahola

## Abstract

Prenatal alcohol exposure (PAE) affects embryonic development, leading to a variable fetal alcohol spectrum disorder (FASD) phenotype with neuronal and dysmorphological defects. To understand the etiology of FASD, we have established a cohort consisting of biological samples at birth and developmental information of prenatally alcohol-exposed and unexposed control children. To explore the phenotypic effects of PAE, we performed neuropsychological, neuropediatric, and dysmorphological assessments on the first 28 PAE children and 52 control children at the age of six.

In the PAE group, 76% (19/25) of the children, who attended a full evaluation, met the criteria for FASD at this age. These diagnoses included five children with fetal alcohol syndrome (FAS), six with partial FAS, seven with alcohol-related neurodevelopmental disorder (ARND), and one with alcohol-related birth defect. In addition to characteristic features of FAS, including short palpebral fissure length, smooth philtrum, and thin vermilion (*P*<0.0001 for each), numerous other anomalies, such as mild midface hypoplasia (*P*<0.0001), were associated significantly with PAE. Unexpectedly, PAE was associated not only with underweight but also with an increased risk of overweight among six-year-olds, prominently among boys.

Neurocognitive assessments indicated lower performance scores and a high prevalence of ADHD symptoms in PAE children compared to controls. Furthermore, additional PAE-associated phenotypic features, such as perturbations in adaptive and social functioning, somatic health issues, as well as reduced postural stability and lower performance in static balance tasks, were observed, which are not included in the diagnostic criteria of FASD. These findings underscore the complexity of FASD diagnosis and the importance of comprehensive phenotypic characterization to improve identification of FASD. This emphasizes the significance of the chosen diagnostic method and may leave some of the tested children without a diagnosis needed for developmental support.

Interestingly, early alcohol exposure, up to the seventh gestational week, resulted in similar rates of dysmorphic features and cognitive scores compared to the longer exposure group. These results, along with several ARND diagnoses in the early PAE group, emphasize the vulnerability of early pregnancy to alcohol exposure, particularly the sensitivity of the nervous system. This is also consistent with the guidelines recommending the use of contraception when consuming alcohol and advising individuals to stop drinking alcohol when planning a pregnancy.

## INTRODUCTION

Prenatal alcohol exposure (PAE) can result in a spectrum of lifelong physical, psychiatric, and cognitive impairments collectively referred to as fetal alcohol spectrum disorders (FASD). These disorders are estimated to affect 2–5% of individuals in the Western world (Lange et al., 2017; Popova et al., 2017; Popova et al., 2023; Roozen et al., 2016). The effects of PAE and the resulting phenotype of affected offspring depend not only on the amount and frequency of exposure, but also on maternal weight, age, concurrent smoking, and genetic susceptibility of both the mother and fetus (Popova et al., 2023). Also, the timing of exposure is critical, and considerable effects of early exposure during the first weeks of embryonic development have been observed (Wallén et al., 2021; Wallén et al., 2025; Lipinski et al., 2012). Owing to these factors, individuals with FASD experience a wide range of neurodevelopmental and somatic disorders, including attention deficit hyperactivity (ADHD), autism spectrum disorder (ASD), schizophrenia, seizures, visual impairments, and middle-ear infections (Attell et al., 2025; Popova et al., 2016). Due to the diverse and age-dependent phenotype, the diagnosis of FASD is challenging.

Certain dysmorphological, neurodevelopmental, and physiological outcomes associated with PAE are encompassed within the diagnostic criteria for FASD (Hoyme et al., 2016; Del Campo et al., 2024). FASD can be classified into four categories according to the Institute of Medicine (IOM) criteria: fetal alcohol syndrome (FAS), partial fetal alcohol syndrome (PFAS), alcohol-related neurodevelopmental disorder (ARND), and alcohol-related birth defect (ARBD). FAS is considered the most severe form of FASD, including restricted growth, functional or structural neuronal defects, and typical facial features such as small eye openings, smooth philtrum, and thin upper lip as its diagnostic criteria (Hoyme et al., 2016). However, the developing nervous system is known to be especially sensitive to the effects of PAE (Guerri, 2002; Wallén et al., 2025) and ARND, including only typical neurodevelopmental impairment, appears to be the most common category of FASD (Wozniak et al., 2019).

To investigate the etiology of FASD, we have established the epiFASD cohort of alcohol-exposed and control newborns. The cohort includes biological samples, as well as pregnancy and birth information collected at delivery, together with follow-up data of the children. To better understand the typical patterns of effects associated with PAE and how they manifest across development, we conducted neuropsychological, neuropediatric, and dysmorphological assessments on 28 PAE children and 52 unexposed controls, analysing differences in their phenotypes at six years of age. This is an important age, considering the future of the affected children, as decisions regarding schooling and necessary support are typically made at this time. Furthermore, to assess the representativeness of the tested children relative to the epiFASD cohort, we analysed information collected from 45 PAE and 47 unexposed control newborns and their mothers, who were also part of the study cohort but did not participate in the tests at the age of six.

## MATERIALS AND METHODS

### The epiFASD cohort

The study examines the phenotypes of children in the epiFASD cohort, which consists of biological samples and follow-up information about newborns with PAE as well as unexposed controls. Informed consent has been obtained from all participants, and we have an ethical approval (HUS/683/2020) as well as permission from Helsinki University Hospital (HUS/706/2025) for the epiFASD cohort. The first 174 children of the cohort born in 2013-2018 turned six years old in 2019-2024, during which the phenotypic assessments were carried out.

Women with substantial alcohol consumption (*n*=73) were recruited in a special outpatient clinic for pregnant women with substance use problems in Helsinki University Hospital, Finland. The amount of maternal alcohol consumption was registered using self-reported information: Alcohol Use Disorders Identification Test (AUDIT) or the number of alcohol units consumed per week (ad) (one unit is 12 g of ethyl alcohol). A 10-item screening tool AUDIT, developed by the World Health Organization, estimates alcohol consumption, drinking behavior, and alcohol-related problems (Babor et al., 2001). Maternal alcohol consumption is presented in three categories according to AUDIT scores or ad (Rehm et al., 2015; Tynjälä et al., 2012): AUDIT scores 1–5 suggest low-risk consumption or < 7 ad cause low risk for morbidity and mortality for non-pregnant women (category 1), AUDIT scores 6–13 suggest hazardous or harmful alcohol consumption or 7–11 ad cause moderate risk for morbidity and mortality for non-pregnant women (category 2), and AUDIT scores 14–40 indicate the likelihood of alcohol dependence (moderate-severe alcohol use disorder) or ≥ 12 ad cause high risk for morbidity and mortality for non-pregnant women (category 3). The timing of maternal alcohol consumption during pregnancy was registered using self-reported information. To avoid specific individual-level data, the timing of consumption is also presented in three categories according to the length of alcohol exposure: gestational weeks (GW) 1–12, GW 1–28, and GW 1–42.

The control mothers (*n*=101), recruited during the years 2014–2016 in Helsinki University Hospital, Finland, were healthy Finnish, Caucasian, who did not use alcohol or smoke during pregnancy according to their self-report.

A total of 80 six-year-old children of the epiFASD cohort who were born in 2013-2018 participated in dysmorphological, neuropediatric, and neuropsychological assessments during 2020-2024: 28 alcohol-exposed (17 females, 11 males) and 52 controls (25 females, 27 males). Although not all 28 alcohol-exposed children completed every assessment, each test included data from at least 25 PAE children. The exposure details were not available for the pediatric neurologist and neuropsychologist during the evaluations. However, it was evident from the medical history of the child if they had been in foster care, adopted, or followed up in the social pediatric unit due to prenatal substance exposure.

Information about maternal age, pre-pregnancy body mass index (BMI), weight gain during pregnancy, and placental weight was collected. Birth measurements, including weight, birth length, and head circumference (HC), were examined using Finnish growth charts (Sankilampi et al., 2013) in which the gestational age (GA) at birth and sex are considered when calculating the standard deviation (*z*-score) of birth measures (SD of birth measures). Furthermore, international standardized body mass index (ISO-BMI), an adjusted version of BMI (childhood BMI was age- and sex-corrected to the adult BMI equivalent) used for children and adolescents aged 2 to 18 years, was used to assess underweight, healthy weight, overweight, and obesity in children based on established adult BMI thresholds (Cole et al., 2000; Cole et al., 2007).

Finally, in the closer examination of the diagnoses, the mothers who consumed alcohol up to GW 7 at maximum were selected in the early PAE subgroup and the rest were included in the longer PAE subgroup.

### Dysmorphological and neuropediatric assessments for six-year-old children

A pediatric neurologist with experience in FASD diagnostics performed a detailed dysmorphology assessment, measurements of height, weight, HC, and palpebral fissure lengths, as well as a full neuropediatric examination, including testing for balance and basic pediatric examination.

#### Dysmorphological assessment

Dysmorphological assessment was performed by detailed inspection, listing any minor anomalies or remarkable personal features, and calculating a dysmorphology score for each individual. The dysmorphology score used was adapted from Hoyme et al. (2016). Adaptation was made by: 1) FAS features (facial gestalt, HC, and growth) included in the diagnostic guidelines were calculated separately, 2) instead of two points, only one point was given for epicanthal folds and anteverted nares, since these features are common in children with Finnish ethnicity (compare Autti-Rämö et al. (2006) and Hoyme et al. (2016)), 3) interpupillary distance was not measured because an ethnicity-specific chart was unavailable, 4) railroad track ears were replaced by any outer ear anomaly, as multiple different types of ear anomalies have been described in FASD and railroad track ears have been shown to be rare among Finnish FASD population (Autti-Rämö et al., 2006), 5) prognathism, hirsutism, and altered palmar creases were classified under “other anomalies” and assigned one point each; however, when prognathism resulted in malocclusion, it was recorded under that category instead, and 6) dental malocclusion and refractive errors requiring glasses by age six were listed as separate categories, as they are commonly recognized in FASD after the 2016 guidelines (Aring et al., 2021; Blanck-Lubarsch et al., 2019; Ludwików et al., 2022; Tsang et al., 2023).

#### Neuropediatric assessment

Neuropediatric assessment was performed using standard neuropediatric examination consisting of:

1. parental/caregiver interview about the developmental and medical history of the child, including the ages at which walking and talking were achieved and any somatic conditions, 2) general observation on behaviour, attention, activity level, eye-contact, interaction, co-operation, speech and understanding, movements, presence of tremor, postures, and mental state, 3) cranial nerves (except for I olfaction), 4) eye examination including pupil reactions, ocular movements, Hirschberg, cover test, and direct ophthalmoscopy, 5) motor examination including tests for Romberg, deep tendon reflexes, Babinski, finger-to-nose, rapid alternating movements, muscular power and tone, range of motion of joints, gait in heel-to-toe, heel and toe walking, hopping with one foot, as well as throwing and catching a ball, 6) static balance was measured in seconds using a stopwatch by having participants stand on one foot with eyes open and closed, and the best of three trials for each foot was recorded, and 7) basic pediatric assessment including stethoscope auscultation of heart and lungs, otoscope examination for ears and throat, visual inspection of skin and hair, as well as manual palpation of neck and abdomen.

### Neuropsychological assessments for six-year-old children

A neuropsychologist performed the following testing:

#### WPPSI-III (WPPSI-III - Wechsler Preschool and Primary Scale of Intelligence - Third Edition)

WPPSI-III was used to assess participants’ general cognitive abilities. The following WPPSI-III were included in analysis: information, vocabulary, word reasoning, block design, matrix reasoning, picture concepts, symbol search, coding, receptive vocabulary, and picture naming. These subtests produce full-scale intelligence quotient (FSIQ), verbal intelligence quotient (VIQ), performance intelligence quotient (PIQ), processing speed quotient (PSQ), and general language composite (GLC) (Luiselli et al., 2013). The standard mean scores of indexes are 100 and the standard deviations are 15. Higher scores indicate better abilities. Used cut-off scores were: FSIQ <70 for intellectual disability range, 70-84 for borderline intellectual functioning, and VIQ ≤ 70 while PIQ >89 for cognitive performance level being typical for developmental language disorder. According to the IOM diagnostic criteria for FASD, the level required cognitive impairment was defined as a score < 78 or −1.5 SD in any analyzed cognitive domain.

#### WISC-IV (Wechsler Intelligence Scale for Children – IV)

Two subtests of WISC-IV measuring working memory were included. These subtests produce the working memory index (WMI) (Grizzle et al., 2011). The standard mean score of index is 100 and the standard deviation is 15. Higher scores indicate better abilities.

#### VINELAND

Adaptive functioning was assessed using Vineland interview for parents (Doll, 1965). Scores were converted to developmental age, and a delay of more than one year was considered significant.

The Following questionnaires were filled out by parent and preschool/kindergarten teacher:

#### ADHD-RS (ADHD Rating Scale IV)

ADHD-RS questionnaire consists of 18 items and measures the frequency of ADHD symptoms (ADHD: Current Care Guidelines 2025; DuPaul et al., 1998). It produces two subscales, which are inattention (9 items) and hyperactivity/impulsivity (9 items). Each item can score 0, 1, 2, or 3 points and the maximum total score for the questionnaire is 54. Higher scores indicate more difficulties. ADHD is likely if the scores exceed the 93rd percentile threshold. ADHD is unlikely if the scores fall below the 85th percentile threshold.

#### SRS (Social Responsiveness Scale)

SRS measures autism spectrum traits in following areas: social awareness, social cognition, social communication, social motivation, and autistic mannerisms including questions about restricted interests and repetitive behavior (Constantino & Gruber, 2005). SRS consists of 65 items. Higher scores indicate more difficulties and T-scores of 60 or more indicates clinically significant difficulties.

#### SDQ (The Strengths and Difficulties Questionnaire)

SDQ is a short screening questionnaire that includes 25 items. It measures emotional symptoms (5 items), conduct problems (5 items), hyperactivity/inattention (5 items), peer relationships problems (5 items), and prosocial behavior (5 items) (Goodman et al., 2001). Each item can score 0, 1, or 2. Higher scores indicated greater difficulties on all scales, except for the prosocial scale, where lower scores indicated more difficulties.

### Evaluation for FASD

Evaluation for FASD was conducted according to the modified 2016 IOM criteria for children over three years of age (Hoyme et al., 2016), which are typically applied in Finland, as the earlier 2005 IOM diagnostic criteria were validated for the Finnish population (Autti-Rämö et al., 2006). The main modification is that when assessing the philtrum and lip, the Four-Digit Code visual guide is used in Finland in place of the IOM guideline, as it offers greater specificity, though with lower sensitivity (Astley et al., 2017; Hemingway et al., 2019). Since Finnish growth charts report z-scores for height, HC, and weight-for-height relative to the mean for age and sex — instead of percentiles (Saari et al., 2011) — the equivalent z-score cut-offs are used in place of the 10th percentile specified in the IOM criteria.

As all children in the PAE group had documented PAE, diagnosing FASD included:

1. Assessment for growth (required in FAS), with used cut-offs of −1.25SD for height and HC as well as −10% for weight-for-height ratio relative to the mean for age and sex.
2. Assessment for facial features of FAS (required for FAS and PFAS), with cut-offs of −1.25SD in the Scandinavian chart (Strömland et al., 1999) for short palpebral fissures and rank 4 or 5 on the Four-Digit Code visual guide for thin upper and smooth philtrum.
3. Assessment for neurobehavioral impairment (required for FAS, PFAS, and ARND), with cut-offs of −1.5 SD (i.e. score 78) for FSIQ, VIQ, PIQ, PSQ, and WMI, as well as 93rd percentile in ADHD-RS score for attention and impulse control, and very high range SDQ score for mood or behavioural regulation.
4. Assessment for malformations (required for ARBD) clinically and by medical history, as no radiological examinations were performed, including congenital heart defects, radioulnar synostosis, vertebral abnormalities, kidney malformations, strabismus, ptosis, retinal vascular anomalies, optic nerve hypoplasia, and hearing loss.

### Statistical analysis

All statistical analyses were performed in IBM SPSS Statistics, version 30.0 (IBM Corp.). All data are presented as the mean ±SD for a normal distribution of variables. Differences between control and PAE groups were calculated, depending on the data distribution and variance, by a parametric two-tailed Student’s *t*-test or a nonparametric Mann-Whitney *U* test. For comparison of proportions, odds ratios (ORs) and their 95% confidence intervals were analysed, along with two-tailed Pearson Chi-Square *P*-values. In cases where an OR could not be estimated due to zero cell counts, a two-tailed Fisher’s exact test was applied. Statistical analyses were performed as described in the figure and table legends or in the relevant sections. Spearman’s rank correlation coefficient, along with two-tailed *P*-values, was used for correlation analysis.

## RESULTS

### Characteristics of the cohort

The general characteristics of epiFASD cohort, including 73 PAE and 101 control newborns, as well as their mothers, were compared (Supplementary Table S1). Control mothers were older, and their gestational weight gain was higher compared to the mothers of PAE children (*P*<0.05, Student’s *t*-test, *P*<0.05, Mann-Whitney *U*, respectively). GA was significantly shorter (*P*<0.01, Mann-Whitney *U*), and weight, length, and HC (SDs) at birth were significantly smaller in PAE children compared to controls (*P*<0.05, *P*<0.05, Mann-Whitney *U*, *P*<0.001, Student’s *t*-test, respectively). At birth, 8/73 (11%) of PAE children had microcephaly (HC< −2 SD) and other malformations were observed at birth in 4/73 (5.5%) children with PAE: three had cleft lip/palate and one had polydactyly. One microcephaly was observed in the control group. Of the children with birth malformations, only one — a child in the PAE group who had microcephaly at birth — participated in the follow-up at age six.

A total of 28 PAE children and 52 unexposed children participated in phenotypic assessments at the age of six. Characteristics of the tested children and information about the pregnancies were analysed (Supplementary Table S1). Also, the representativeness of the tested children relative to the cohort was assessed by comparing the pregnancies of tested and untested PAE children. Among the cohort, no significant differences in the amount or timing of maternal alcohol consumption between tested and untested pregnancies were observed (Figure 1a). Alcohol consumption by mothers was substantial, particularly during the first trimester (Figure 1b-d). Furthermore, smoking was prevalent among mothers in the PAE group, 62/73 (84.9%) of infants had at least early prenatal smoking exposure and 43/73 (58.9%) had prenatal smoking exposure throughout gestation. The groups did not differ from each other in this regard (82.2% of untested and 89.3% of tested were smoking-exposed). Moreover, maternal medication or drug use did not differ significantly between the groups (Supplementary Table S1).

**Figure 1.**
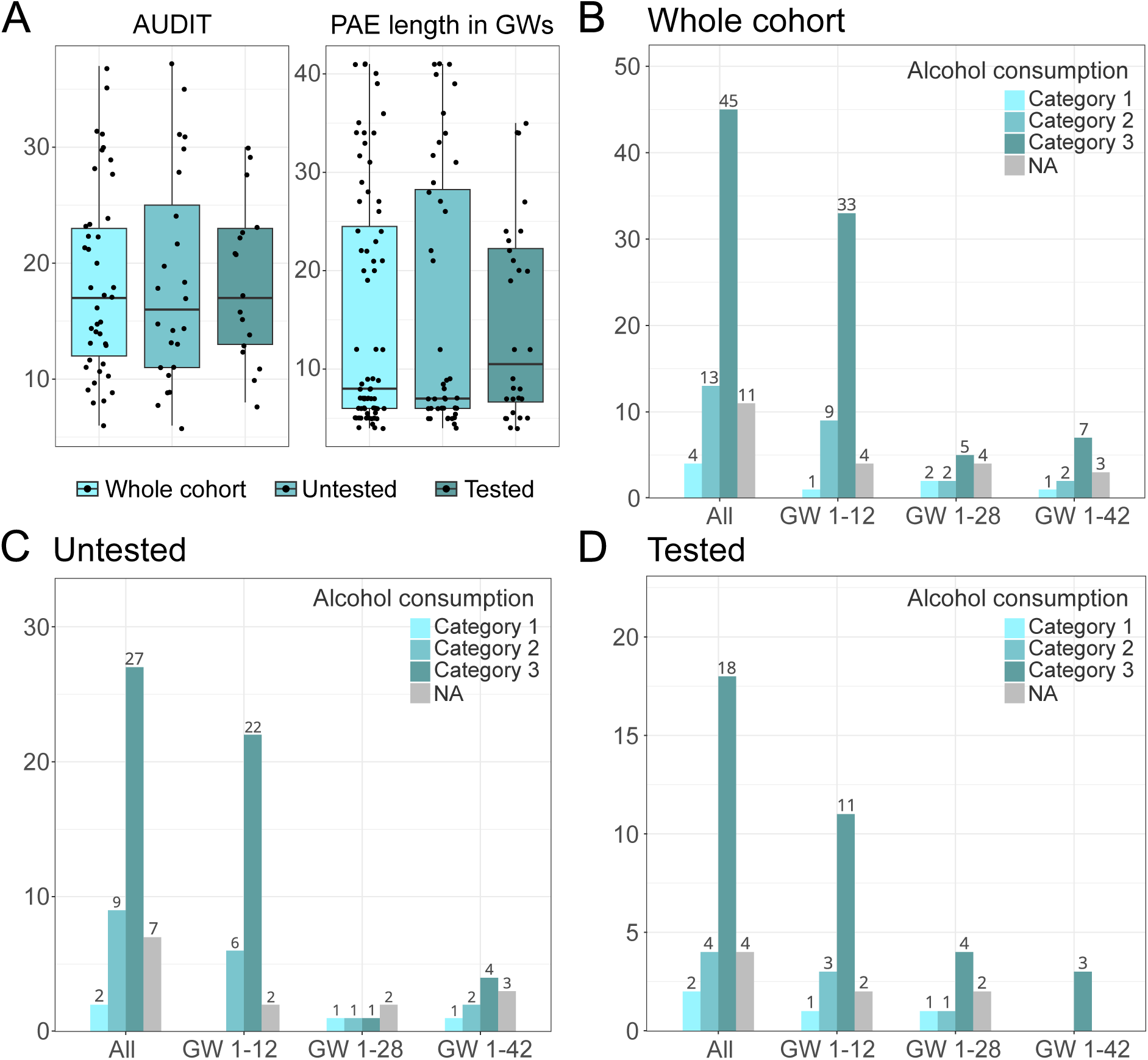
Amount and timing of maternal alcohol consumption in the whole cohort as well as in the subgroups of untested and tested PAE children. a) AUDIT scores and PAE length in GWs (mean ±SD) in the original cohort (*n*=73), untested (*n*=45) and tested (*n*=28) groups. No significant differences in the amount or timing of maternal alcohol consumption between groups were observed. Amount and timing of maternal alcohol consumption in categories in b) the whole cohort, c) untested, and d) tested subgroup. Categories for maternal alcohol consumption: AUDIT scores 1–5 suggest low-risk consumption or < 7 alcohol units consumed per week (ad) cause low risk for morbidity and mortality for non-pregnant women (category 1), AUDIT scores 6–13 suggest hazardous or harmful alcohol consumption or 7–11 ad cause moderate risk for morbidity and mortality for non-pregnant women (category 2), and AUDIT scores 14–40 indicate the likelihood of alcohol dependence (moderate-severe alcohol use disorder) or ≥ 12 ad cause high risk for morbidity and mortality for non-pregnant women (category 3). Categories for GW: GW 1–12, GW 1–28, and GW 1–42. NA: not applicable.

The GA of the tested controls was significantly longer compared to the untested controls (*P*<0.0001, Student’s *t*-test). When anthropometrics was compared between untested PAE and control children, as well as tested PAE and control children, in both PAE groups, significantly smaller HCs (SD) were observed (*P*<0.01, *P*<0.05, respectively, Student’s *t*-test). Among untested PAE children, there was a higher number of small HCs (HC ≤ −1.25 SD) compared to the tested PAE (Supplementary Table S1).

### Characteristics of the tested six-years-olds

The general characteristics of tested six-year-olds and their parents were compared (Table 1). Control parents had higher education compared to the parents of PAE children. Four families in the PAE group (16.7%) and one family in the control group (1.9%) had a history of ASD. In the PAE group, there were four adopted children and three in foster care.

**Table 1.** General characteristics of tested six-years-old PAE and control children. Differences in weight, length, and HC were calculated using both anthropometric measures and the SDs (z-scores) of measures based on Finnish growth charts (Sankilampi et al., 2013). *P*-values were calculated using 1) Student’s t-test (equal variances not assumed, two-tailed *P*-value), 2) Student’s t-test (equal variances assumed, two-tailed *P*-value), 3) Mann-Whitney U (exact two-tailed *P*-value), 4) OR (intervals, Pearson Chi-Square two-tailed *P*-value), and 5) Fisher’s exact test (two-tailed *P*-value). BMI: body mass index, GA: gestational age, HC: head circumference, Weight-for-height: difference in percents from mean for age and sex. ISO-BMI: childhood BMI age- and sex-corrected to the adult BMI equivalent, underweight: ISO-BMI<18, overweight: ISO-BMI >25.

|  |  | Control<br>(n = 52)<br>mean (±SD) | PAE<br>(n = 28)<br>mean (±SD) | P-value or<br>OR (95% CI, P-value) |
| --- | --- | --- | --- | --- |
| **Maternal characteristics** |  |  |  |  |
| Age | (years) | 31.3 (±4.5) | 29.4 (±6.2) | 0.159<sup>(1)</sup> |
| Pre-pregnancy BMI | (kg/m<sup>2</sup>) | 22.8 (±3.2) n=52 | 25.5 (±6.2) n=22 | 0.191<sup>(3)</sup> |
| Gestational weight gain | (kg) | 14.9 (±3.7) n=49 | 15.1 (±5.8) n=19 | 0.899<sup>(1)</sup> |
| Parity | (% of firstborns) | 30 (57.7%) | 20 (71.4%) | 1.8 (0.7-4.9, P=0.333)<sup>(4)</sup> |
| **At birth** |  |  |  |  |
| Sex | (males/females) n (%) | 27/25 (51.9/48.1%) | 11/17 (39.3/60.7%) | 1.7 (0.7-4.2, P=0.350)<sup>(4)</sup> |
| GA | (days) | 286.5 (±7.1) | 276.4 (±14.9) | <0.01<sup>(1)</sup> |
| GA < 38 weeks | n (%) | 0 (0%) | 5 (17.9%) | <0.01<sup>(5)</sup> |
| Birth weight | (g) | 3670 (±415) | 3240 (±526) | <0.001<sup>(2)</sup> |
|  | (SD) | -0.09 (±1.0) | -0.48 (±0.9) | 0.088<sup>(2)</sup> |
| Birth weight ≤ -1.25 SD | n (%) | 7 (13.5%) | 4 (14.3%) | 1.1 (0.3-4.0, P=1)<sup>(4)</sup> |
| Birth length | (cm) | 50.4 (±1.6) | 48.9 (±2.2) | <0.001<sup>(3)</sup> |
|  | (SD) | -0.13 (±0.90) | -0.91 (±0.96) | 0.062<sup>(2)</sup> |
| Birth length ≤ -1.25 SD | n (%) | 6 (11.5%) | 8 (28.6%) | 3.1 (0.9-10.0, P=0.070)<sup>(4)</sup> |
| HC | (cm) | 35.6 (±1.2) | 34.3 (±2.6) | <0.001<sup>(3)</sup> |
|  | (SD) | 0.22 (±0.87) | -0.24 (±1.07) | <0.05<sup>(2)</sup> |
| HC ≤ -1.25 SD | n (%) | 3 (5.8%) | 4 (14.3%) | 2.7 (0.6-13.1, P=0.232)<sup>(4)</sup> |
| **At 6 years of age** |  | n=50 | n=25 |  |
| Weight | (kg) | 22.3 (±2.9) | 24.6 (±6.9) | 0.559<sup>(3)</sup> |
|  | (Weight-for-height) | 0.54 (±8.9) | 7.48 (±21.17) | 0.653<sup>(3)</sup> |
|  | (ISO-BMI) | 21.7 (±3.4) | 26.2 (±11.8) | 0.385<sup>(3)</sup> |
| Underweight | n (%) | 5 (10%) | 3 (12%) | 1.2 (0.3-5.6, P=1)<sup>(4)</sup> |
| Ever been underweight | n (%) | 5 (10%) | 7 (28%) | 2.8 (0.8-10.5, P=0.164)<sup>(4)</sup> |
| Overweight | n (%) | 4 (8%) | 7 (28%) | 4.4 (1.2-17.2, P<0.05)<sup>(4)</sup> |
| Height | (cm) | 118.6 (±4.6) | 119.0 (±4.9) | 0.739<sup>(2)</sup> |
|  | (SD) | -0.03 (±1.42) | 0.02 (±1.09) | 0.831<sup>(2)</sup> |
| Height ≤ -1.25 SD | n (%) | 4 (8%) | 3 (12%) | 1.6 (0.3-7.6, P=0.680)<sup>(4)</sup> |
| HC | (cm) | 52.6 (±1.3) | 52.0 (±1.6) | 0.085<sup>(3)</sup> |
|  | (SD) | 0.03 (±1.0) | -0.43 (±1.3) | 0.084<sup>(2)</sup> |
| HC ≤ -1.25 SD | n (%) | 3 (6%) n=49 | 7 (28%) | 6.0 (1.4-26.2, P<0.05)<sup>(4)</sup> |
| **Family background** |  |  |  |  |
| **Education of the higher-educated parent** |  | n=51 | n=24 |  |
| Basic education (9 years or less) | n (%) | 0 (0%) | 3 (12.5%) | <0.05<sup>(5)</sup> |
| Completed secondary education (12 years) | n (%) | 7 (13.7%) | 10 (41.7%) | 4.6 (1.5-14.3, P<0.01)<sup>(4)</sup> |
| Post-secondary or tertiary education (>12 years) | n (%) | 44 (86.3%) | 11 (45.8%) | 0.1 (0.0-0.4, P<0.001)<sup>(4)</sup> |
| **Education of the other parent** |  | n=43 | n=16 |  |
| Basic education (9 years or less) | n (%) | 0 (%) | 4 (25%) | <0.01<sup>(5)</sup> |
| Completed secondary education (12 years) | n (%) | 10 (23.3%) | 5 (31.3%) | 1.5 (0.4-5.4, P=0.738)<sup>(4)</sup> |
| Post-secondary or tertiary education (>12 years) | n (%) | 33 (76.7%) | 7 (43.8%) | 0.2 (0.1-0.8, P<0.05)<sup>(4)</sup> |
| **Family history** |  | n=52 | n=25 |  |
| Attention difficulties | n (%) | 7 (13.5%) | 8 (32%) | 3.0 (1.0-9.6, P=0.069) |
| ASD | n (%) | 1 (1.9%) | 4 (16.7%) n=24 | 10.2 (1.1-96.9, P<0.05) |
| Language difficulties | n (%) | 12 (23.1%) | 6 (25%) n=24 | 1.1 (0.4-3.4, P=1) |
| Other learning difficulties | n (%) | 2 (3.8%) | 3 (12.5%) n=24 | 3.6 (0.6-23.0, P=0.318) |
|  |  | n=52 | n=25 |  |
| Bilingual/multilingual | n (%) | 7 (13.5%) | 4 (16.7%) n=24 | 1.3 (0.4-4.9, P=0.734)<sup>(4)</sup> |
| Adoption | n (%) | 0 (0%) | 4 (16%) | <0.01<sup>(5)</sup> |
| Foster care | n (%) | 0 (0%) | 3 (12%) | <0.05<sup>(5)</sup> |

Even though PAE newborns had a trend of lower birth weight at birth, overweight (ISO-BMI >25) was more common in the PAE group compared to controls at the age of six (OR 4.4, 95% CI 1.2-17.2, *P*<0.05) (Figure 2, Table 1). Due to a few significantly overweight children, the mean weight in the PAE group was actually higher than in the control group. Interestingly, only a minority of boys (3/10) in the PAE group had weight within normal limits relative to their age and height; two (20%) were underweight, and five (50%) were overweight (Supplementary Figure S1). In control boys, only three (10.7%) were underweight and only two (7.1%) were overweight. Furthermore, in addition to the smaller HCs (SD) at birth (*P*<0.05, Student’s *t*-test), smaller HCs (≤ −1.25 SD) were more common in the PAE group than in the control group at the age of six (OR 6.0, 95% CI 1.4-26.2, *P*<0.05).

**Figure 2.**
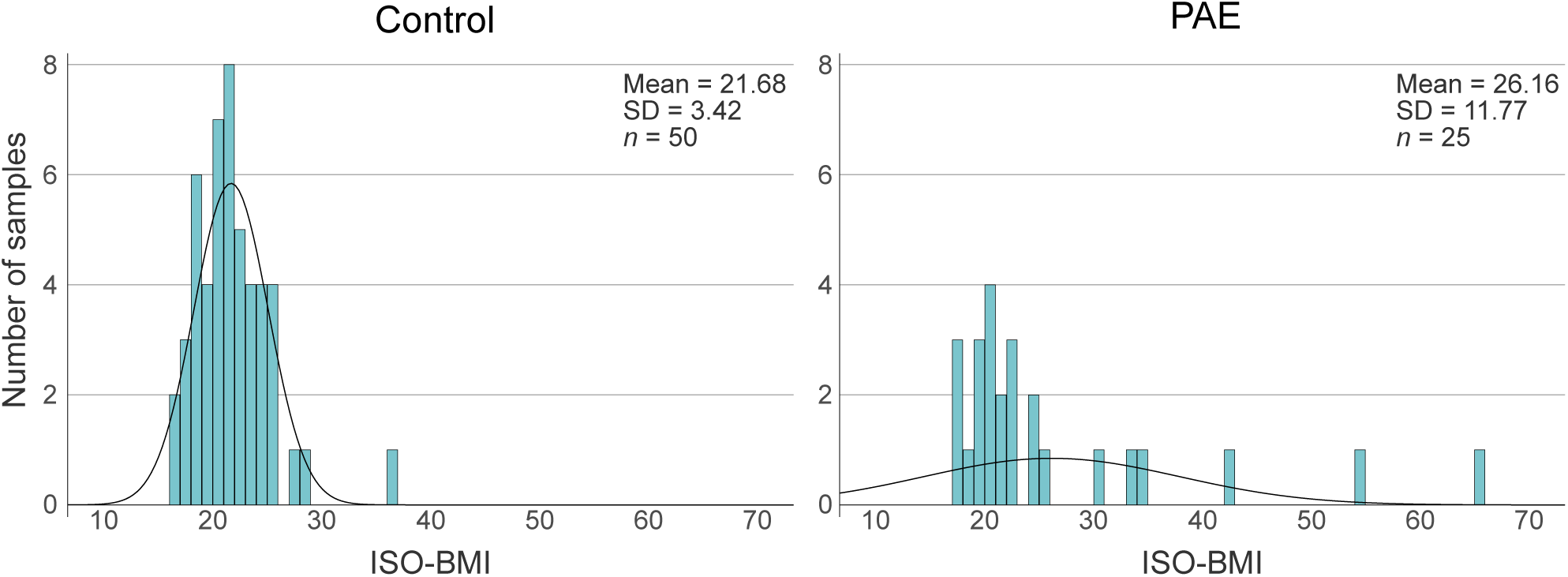
Assessment of six-year-olds weight status by International Standardized Body Mass Index (ISO-BMI). A higher number of overweight (ISO-BMI>25) children was observed in the PAE group compared to controls. ISO-BMI: childhood BMI age- and sex-corrected to the adult BMI equivalent.

### Dysmorphological phenotypes at age six

Children with PAE had significantly more dysmorphic features than controls, with both total FAS dysmorphology scores and all dysmorphology scores notably higher in the PAE group (*P*<0.0001, Mann-Whitney *U*) (Table 2). In addition to those associated with FAS dysmorphology, such as short palpebral fissure length (PFL), smooth philtrum, and thin vermilion, there were numerous other minor anomalies. Mild midface hypoplasia showed a particularly strong association with PAE (OR 40.3, 95% CI 9.4-173.0, *P*<0.0001).

**Table 2.** FAS dysmorphology scores and other dysmorphology scores of six-year-old PAE and control children. *P*-values were calculated using 1) OR (intervals, Pearson Chi-Square two-tailed *P*-value), 2) Mann-Whitney U (exact two-tailed *P*-value), and 3) Fisher’s exact test (two-tailed *P*-value). PFL: palpebral fissure length.

| Feature | Score | Control<br>(*n* = 50)<br>*n* (%) | PAE<br>(*n* = 25)<br>*n* (%) | *P*-value or<br>OR (95% CI, *P*-value) |
| --- | --- | --- | --- | --- |
| **FAS dysmorphology** |  |  |  |  |
| Head circumference < -1.25 SD | 3 | 3 (6.1%) *n*=49 | 7 (28%) | 5.9 (1.4-25.7, *P*<0.05)<sup>(1)</sup> |
| Weight/height ratio < -10% of mean | 1 | 4 (8%) | 4 (16%) | 2.2 (0.5-9.6, *P*=0.429)<sup>(1)</sup> |
| Height < -1.25 SD | 2 | 4 (8%) | 3 (12%) | 1.5 (0.3-7.6, *P*=0.680)<sup>(1)</sup> |
| Short PFL < -1.25 | 3 | 2 (4.2%) *n*=48 | 15 (60%) | 34.5 (6.8-175.4, *P*<0.0001)<sup>(1)</sup> |
| Smooth philtrum grade IV or V | 3 | 4 (8%) | 13 (52%) | 12.5 (3.4-45.2, *P*<0.0001)<sup>(1)</sup> |
| Thin vermilion grade IV or V | 3 | 2 (4%) | 11 (44%) | 18.9 (3.7-95.3, *P*<0.0001)<sup>(1)</sup> |
| **Total FAS score Mean (SD)** | Max 15 | 0.86 (±1.44) | 6.08 (±3.46) | <0.0001<sup>(2)</sup> |
| **Other minor anomalies** |  |  |  |  |
| Hypoplastic midface | 2 | 3 (6%) | 18 (72%) | 40.3 (9.4-173.0, *P*<0.0001)<sup>(1)</sup> |
| Long philtrum | 2 | 3 (6%) | 8 (32%) | 7.4 (1.8-31.1, *P*<0.01)<sup>(1)</sup> |
| Flat nasal bridge | 2 | 4 (8%) | 7 (28%) | 4.5 (1.2-17.1, *P*<0.05)<sup>(1)</sup> |
| Camptodactyly | 2 | 0 (%) | 1 (4%) | 0.324<sup>(3)</sup> |
| Ptosis | 2 | 2 (4%) | 1 (4%) | 1 (0.1-11.6, *P*=1)<sup>(1)</sup> |
| 5th finger clinodactyly | 2 | 7 (14%) | 7 (28%) | 2.4 (0.7-7.8, *P*=0.208)<sup>(1)</sup> |
| Strabismus | 1 | 2 (4%) | 5 (20%) | 4.6 (0.8-27.0, *P*=0.170)<sup>(1)</sup> |
| Refraction disorder, glasses at 6 yrs | 1 | 3 (6%) | 2 (8%) | 1.4 (0.2-8.7, *P*=1)<sup>(1)</sup> |
| Epicanthal folds | 1 | 14 (28%) | 14 (56%) | 3.3 (1.2-8.9, *P*<0.05)<sup>(1)</sup> |
| Anteverted nares | 1 | 17 (34%) | 15 (60%) | 2.9 (1.1-7.9, *P*<0.05)<sup>(1)</sup> |
| Limited elbow supination | 1 | 1 (2%) | 4 (16%) | 9.3 (1.0-88.6, *P*<0.05)<sup>(1)</sup> |
| Outer ear anomaly | 1 | 8 (16%) | 9 (36%) | 3.0 (1.0-9.0, *P*=0.078)<sup>(1)</sup> |
| Dental malocclusion | 1 | 4 (8%) | 4 (16%) | 2.2 (0.5-9.6, *P*=0.429)<sup>(1)</sup> |
| Hypoplastic fingernails | 1 | 0 (%) | 1 (4%) | 0.333<sup>(3)</sup> |
| Any other minor anomaly | 1 each | 10 (20%) | 8 (32%) | 1.9 (0.6-6.0, *P*=0.390)<sup>(1)</sup> |
| **All dysmorphology score Mean (SD)** | Max 25 | 3.06 (±2.53) | 12.28 (±4.51) | <0.0001<sup>(2)</sup> |

Moreover, correlations between FAS and all dysmorphology scores as well as maternal alcohol consumption (AUDIT scores), length of the PAE in weeks, and anthropometrics at birth and at the age of six were examined (Supplementary Table S8). AUDIT scores correlated significantly with all dysmorphology scores (*r*=0.554, *P*<0.05) and length of the PAE with FAS dysmorphology scores (*r*=0.412, *P*<0.05). Also, negative correlations between FAS dysmorphology scores and both HC and weight (SD) at birth (*r*=-0.303, *P*<0.01; *r*=-0.252, *P*<0.05, respectively) and HC (SD) at the age of six (*r*=-0.351, *P*<0.01) were observed. All dysmorphology scores correlated negatively with HC and weight (SD) at birth (*r*=-0.412, *P*<0.05; *r*=-0.241, *P*<0.05).

### Neuropediatric examination

The age at which the tested children began walking and talking in phrases was delayed more often in the PAE group compared to controls, according to parental reports. In the PAE group, 4/23 (17%) children and in the control group, 3/50 (6%) children began walking after 15 months of age. However, the difference was not statistically significant (Supplementary Table S2). Furthermore, a significantly higher number of PAE children 7/20 (35%) started to talk in phrases after two years of age compared to controls 6/49 (12%) (OR 3.9, 95% CI 1.1-13.5, *P*<0.05).

Static balance (seconds standing on one foot), both eyes open and closed, was measured, and it was significantly weaker in the PAE group compared to controls (*P*<0.05, *P*=0.01, respectively, Mann-Whitney *U*) (Figure 3a). Weak balance, defined as having the sum of both feet static balance less than 20 seconds eyes open or less than 5 seconds eyes closed, was more common in the PAE group, 6/25 (24%), compared to controls 4/50 (8%). However, a statistically significant difference was not reached (Supplementary Table S2).

**Figure 3.**
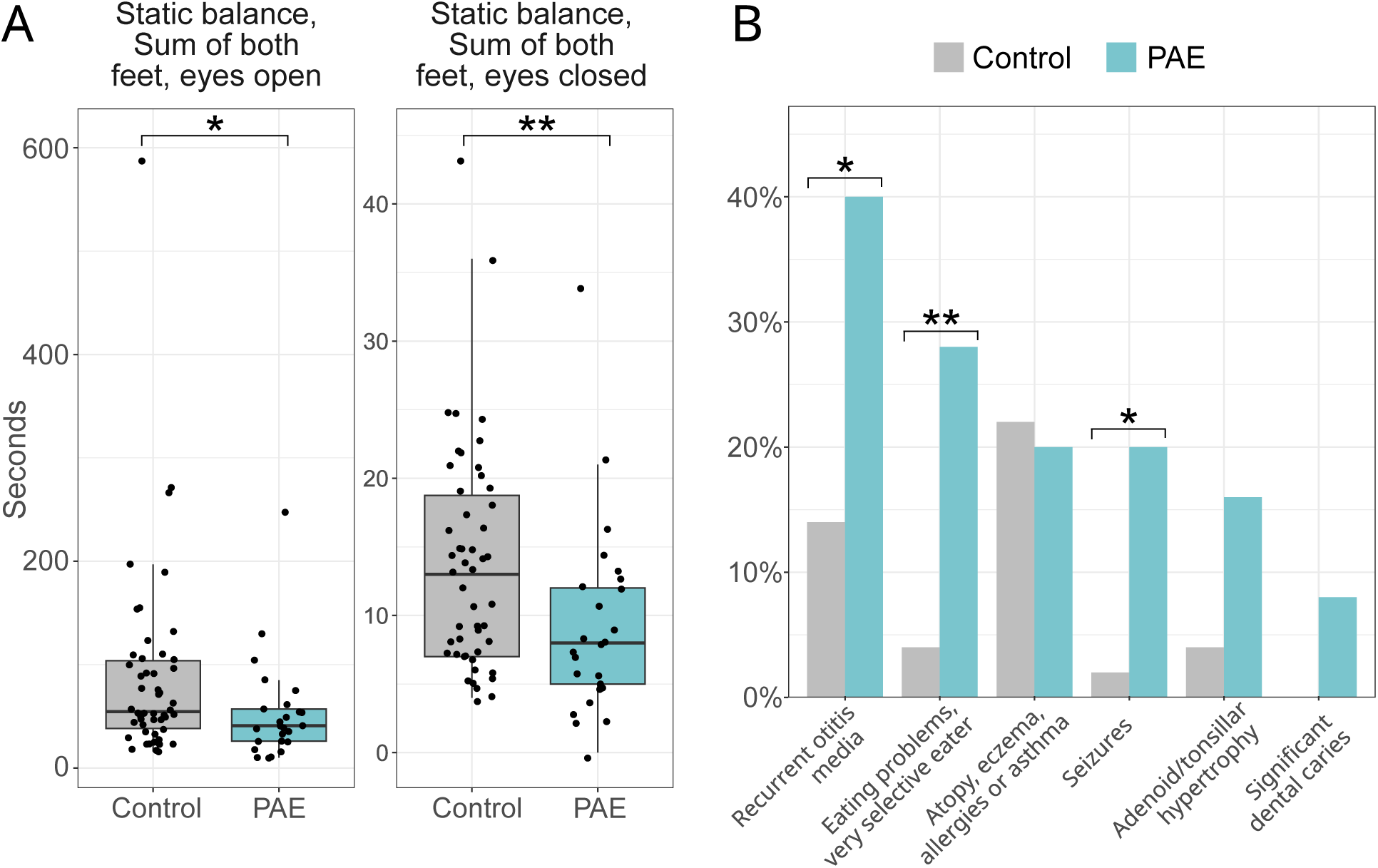
Static balance and somatic conditions of six-year-old PAE and control children. a) Static balance, which was measured by standing on one foot with eyes open and closed (best of three tries for each foot), was weaker in the PAE group compared to controls. b) Increased number of somatic conditions were observed in the PAE group (eyesight problems are included in dysmorphology). * *P*-value <0.05. ** *P*-value <0.01.

Furthermore, children with PAE had increased risks to somatic conditions, such as recurrent middle ear infections (OR 4.1, 95% CI 1.3-12.7, *P*<0.05), very selective eating (OR 9.3, 95% CI 1.8-49.2, *P*<0.01), and history of seizures (OR 12.3, 95% CI 1.4-111.6, *P*<0.05)) (Figure 3b, Supplementary Table S3).

### Neuropsychological examination

#### Cognitive functioning

Children with PAE showed lower average performance on cognitive tests compared to the control group (Figure 4, Supplementary Table S4). FSIQ, VIQ, PIQ, and WMI scores were significantly lower in PAE children compared to controls (*P*<0.05, *P*<0.05, *P*<0.01, and *P*<0.01, respectively, Student’s *t*-test). In the PAE group, global developmental delay, corresponding to borderline intellectual functioning with FSIQ scores 70-84 as well as both VIQ and PIQ <90, was observed in six (24%) children, whereas two (8.3%) performed at the level of mild intellectual disability (FSIQ 50-69) and two (8.3%) performed at the level typical for developmental language disorder (VIQ ≤70, PIQ >89). None in the control group had global developmental delay, but four (7.7%) exhibited difficulties in language performance. FASD diagnostic criteria for cognitive impairment were met in 10/25 (40%) of PAE children, who in neuropsychological testing performed in one or more tested domains (FSIQ, VIQ, PIQ, WMI, and/or PSQ) below 1.5 SD below the mean (<78 IQ points).

**Figure 4.**
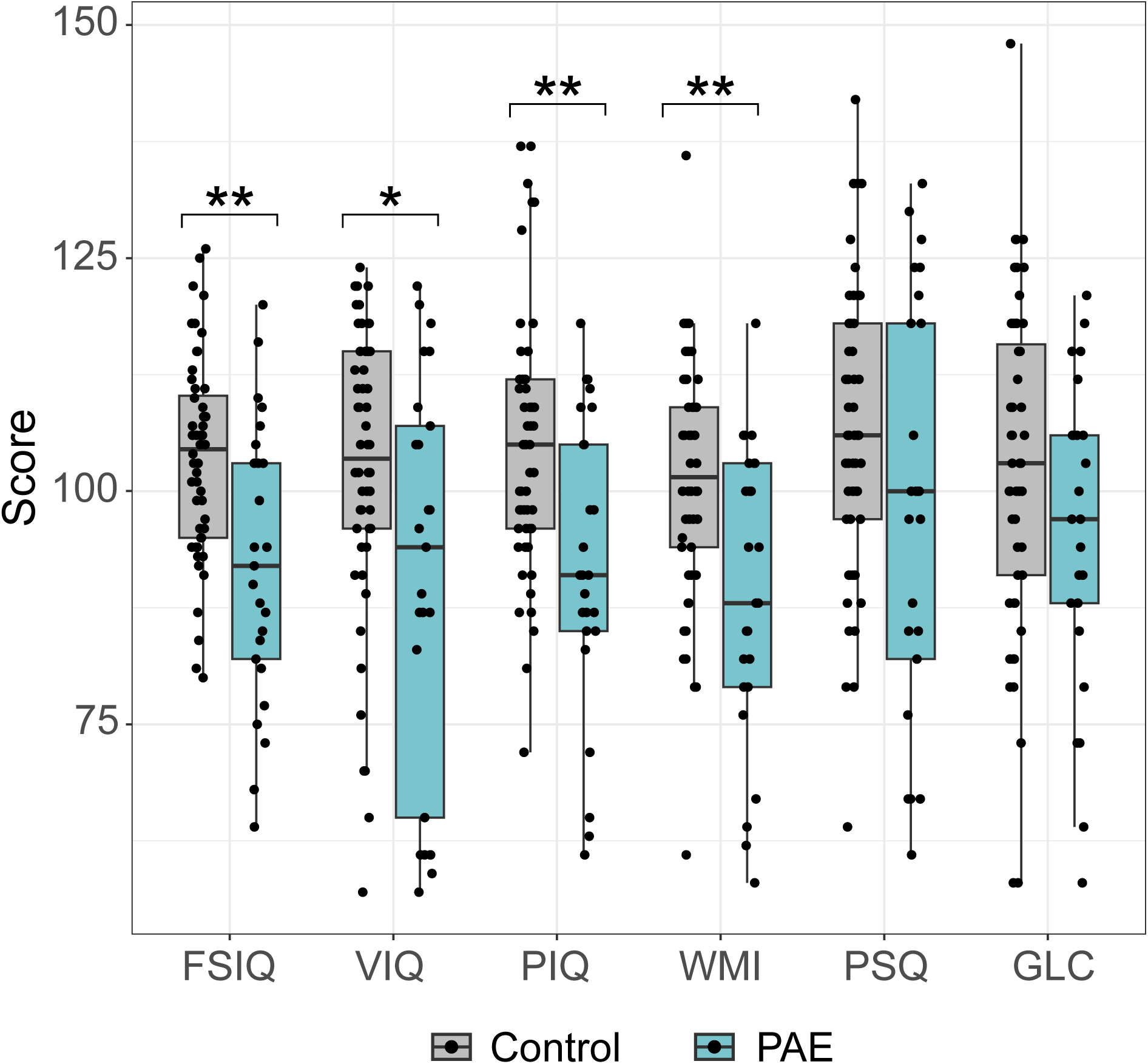
Performance of six-year-old PAE and control children in cognitive tests. FSIQ, VIQ, PIQ, and WMI scores were significantly lower in PAE children compared to controls. FSIQ: Full Scale Intelligent Quotient (IQ), VIQ: Verbal IQ, PIQ: Performance IQ, WMI: Working Memory Index, PSQ: Processing Speed Quotient, GLC: General Language Composite. * *P*-value <0.05. ** *P*-value <0.01.

Correlations between cognitive test scores and AUDIT scores, length of the PAE, anthropometrics, as well as FAS and all dysmorphology scores were examined (Supplementary Table S8). Significant negative moderate correlations between FAS dysmorphology scores and FSIQ (*r*=-0.354, *P*<0.01), VIQ (*r*=-0.241, *P*<0.05), PIQ (*r*=-0.345, *P*<0.01), WMI (*r*=-0.302, *P*<0.01), and PSQ (*r*=-0.232, *P*<0.05) were observed. Also, all dysmorphology scores correlated negatively with FSIQ (*r*=-0.334, *P*<0.01), VIQ (*r*=-0.230, *P*<0.05), PIQ (*r*=-0.290, *P*<0.05), and WMI (*r*=-0.304, *P*<0.01). Also, HC (SD) at the age of six correlated positively with PIQ (*r*=0.261, *P*<0.05).

#### Adaptive functioning

The adaptive functioning was scored, and the difference between socioemotional developmental age and chronological age in daily life functioning was assessed using the Vineland interview with parents (Supplementary Table S5). The total score as well as the difference between socioemotional age and chronological age were significantly lower in PAE children compared to the control group (*P*<0.001, Mann-Whitney *U*). Developmental delays of over one year were more common in the PAE group (10/26; 38.5%) compared to controls (1/48; 2.1%) (OR 29.4, 95% CI 3.5-247.8, *P*<0.0001). Also, communication, general self-help ability, and self-help dressing ability scores were lower in PAE children (*P*<0.05 in all, Mann-Whitney *U*).

#### Neurobehavioral and social functioning

ADHD symptoms, assessed using ADHD-RS and SDQ (hyperactivity category) questionnaires completed by parents and teachers, were significantly more prevalent in the PAE group in all categories: total, inattention, and hyperactivity/impulsivity (Figure 5, Supplementary Table S6). Based on ADHD-RS being >93^rd^ percentile in any category (total, inattention, hyperactivity/impulsivity) or SDQ hyperactivity score in “very high” category, along with presenting significant attention problems and/or hyperactivity/impulsivity in neuropsychological and neuropediatric assessments, clinically significant ADHD symptoms were observed in 11 (44.4%) children with PAE, compared to 2 (3.8%) in the control group.

**Figure 5.**
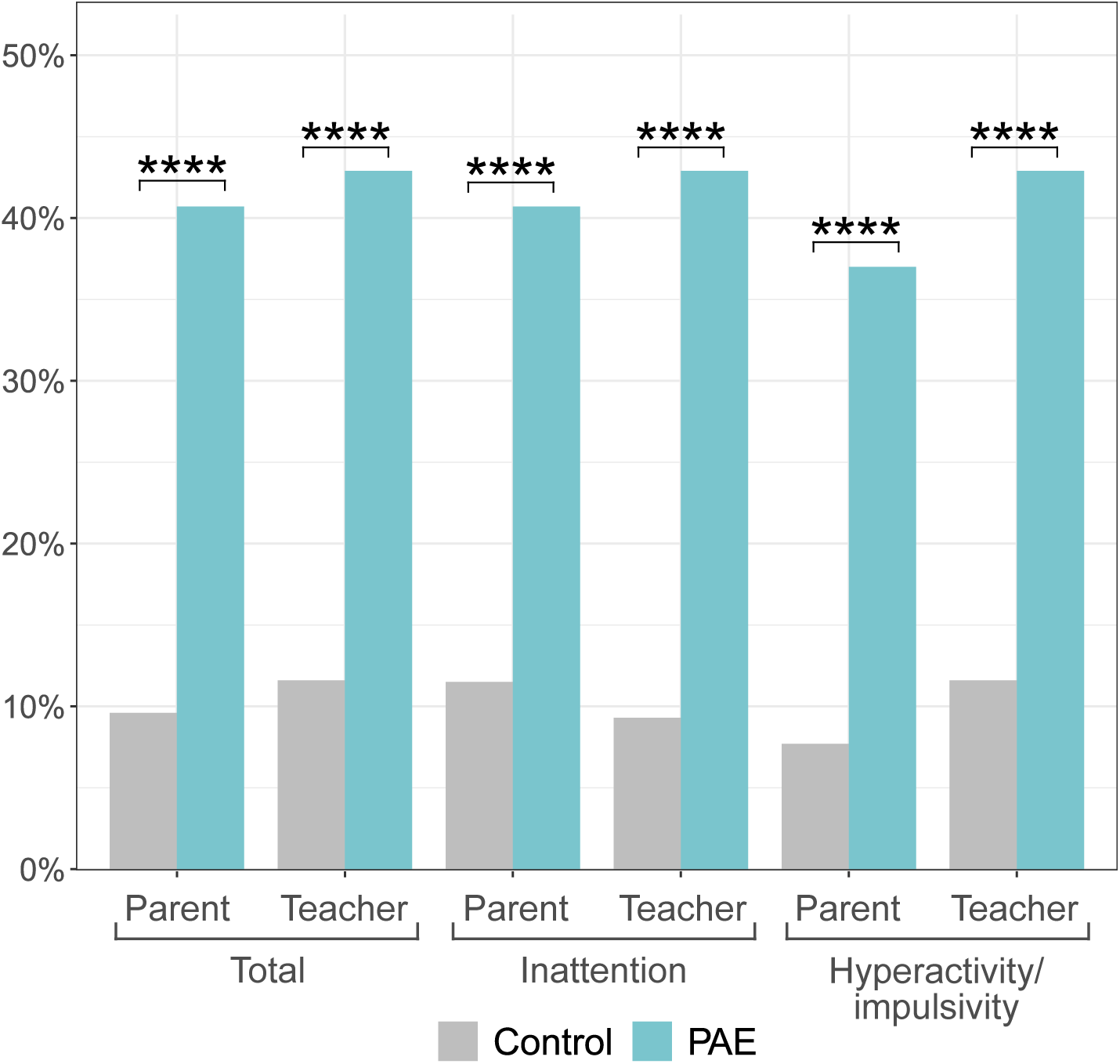
Percentages of PAE and control children having elevated (>85^th^ percentile) ADHD-RS scores reported by parents and teachers in categories total, inattention, and hyperactivity/impulsivity at the age of six. **** *P*-value <0.0001.

According to SDQ questionnaire problem score being in the “very high” range in any of the following categories: total, emotional, and conduct, in addition to aforementioned ADHD symptoms, a total of 14/24 (58.3%) of PAE children met the FASD criteria for neurobehavioral impairment in at least one domain: self-regulation, behavioral regulation, mood regulation, attention deficit, or impulse control.

Even though social functioning problems are not included in the used FASD criteria, based on the SRS questionnaire scores and SDQ categories of prosocial behavior and peer difficulties, both parents and teachers of PAE children reported greater social functioning difficulties than those of control children (Supplementary Table S7). In both groups, parents noted slightly more problems than teachers.

When correlations between ADHD-RS scores and AUDIT scores, length of the PAE, anthropometrics as well as FAS and all dysmorphology scores were examined, significant positive correlation between AUDIT scores and inattention reported by parents (*r*=0.614, *P*<0.05) as well as between ADHD-RS scores reported by parents and teachers and both FAS dysmorphology scores (*r*=0.439, *P*<0.001; *r*=0.487, *P*<0.001, respectively) and all dysmorphology scores (*r*=0.328, *P*<0.01; *r*=-0.354, *P*<0.01, respectively) were observed (Supplementary Table S8). Also, a negative moderate correlation was detected between ADHD-RS scores reported by parents and teachers and HC (SD) at the age of six (*r*=-0.262, *P*<0.05; *r*=-0.345, *P*<0.01, respectively).

Additionally, when correlations between total problem scores of SRS and SDQ questionnaires reported by parents and teachers as well as previously mentioned variables were calculated, a positive correlation was observed with AUDIT and SRS scores reported by parents (*r*=0.547, *P*<0.05) as well as both SRS and SDQ scores and FAS dysmorphology scores, all dysmorphology scores, and HC (SD) at the age of six (Supplementary Table S8).

### FASD diagnoses

According to dysmorphological and neuropsychological assessments, 19 (76%) out of 25 PAE children met criteria for FASD diagnoses at the age of six (Table 3). A total of five FAS, six PFAS, seven ARND, and one ARBD diagnoses were observed. Of those six children who did not meet FASD criteria, two showed facial features characteristic of FAS, but no significant cognitive, behavioral, or growth impairments at this age. Furthermore, among those six children, two had impaired adaptive functioning, but this is not included in the used FASD criteria.

**Table 3.** FASD diagnoses and other FASD-associated features of tested six-year-old PAE children. All FASD category consists of FAS, PFAS, ARND, and one ARBD diagnoses. *cognitive impairment = in at least one domain <-1.5 SD or <78 points, one child did not participate in neuropsychological evaluation. **total SRS T-score >59, parent’s SDQ peer difficulties >4, teacher’s SDQ peer difficulties >5, parent’s SDQ prosocial <6, or teacher’s SDQ prosocial <4. *** according to the Vineland interview, the individual’s socioemotional developmental age was more than one year below the chronological age. HC: head circumference, FSIQ: Full Scale Intelligent Quotient, EEG: electroencephalography.

|  |  | FAS<br>(n = 5) | PFAS<br>(n = 6) | ARND<br>(n = 7) | All FASD<br>(n = 18 + ARBD) | No FASD<br>diagnosis yet<br>(n = 6) |
| --- | --- | --- | --- | --- | --- | --- |
| **FASD diagnostic criteria** |  |  |  |  |  |  |
| FAS dysmorphic score | mean (±SD) | 10.4 (±1.9) | 7.5 (±1.6) | 4.0 (±3.1) | 6.7 (±3.5) | 4 (±2.5) |
| Growth impairment | n (%) | 5 (100%) | 0 (0%) | 3 (42.9%) | 9 (47.4%) | 0 (0%) |
| HC < -1.2SD | n (%) | 4 (80%) | 2 (33.3%) | 1 (14.2%) | 7 (36.8%) | 0 (0%) |
| Cognitive impairment* | n (%) | 2 (50%) n=4 | 3 (50%) | 5 (71.4%) | 10 (52.6%) | 0 (0%) |
| FSIQ | mean (±SD) | 87.0 (±9.1) | 87.3 (±17.5) | 86.4 (±11.9) | 88.5 (±14.2) | 106.5 (±7.4) |
| Behavioral impairment | n (%) | 5 (100%) | 4 (66.7%) | 5 (71.4%) | 14 (73.7%) | 0 (0%) |
| ADHD | n (%) | 4 (80%) | 3 (50%) | 4 (57.1%) | 11 (57.9%) | 0 (0%) |
| ADHD-RS total score (parent) | mean (±SD) | 33.8 (±10.1) | 20.3 (±6.1) | 24.3 (±12.0) | 23.8 (±11.2) | 10.3 (±5.8) |
| **Other FASD-associated features** |  |  |  |  |  |  |
| History of any seizures, EEG performed | n (%) | 2 (40%) | 1 (16.7%) | 0 (0%) | 5 (26.3%) | 0 (0%) |
| Total dysmorphic score | mean (±SD) | 17.8 (±3.8) | 13.3 (±2.9) | 9.7 (±4.0) | 13.1 (±4.6) | 9.7 (±3.1) |
| Social impairment** | n (%) | 2 (40%) | 4 (66.7%) | 5 (71.4%) | 15 (79.0%) | 0 (0%) |
| Adaptive impairment*** | n (%) | 4 (80%) | 1 (16.7%) | 2 (28.6%) | 7 (36.8%) | 2 (33.3%) |
| Coordination disorder (F82) | n (%) | 1 (20%) | 2 (33.3%) | 1 (14.2%) | 4 (21.1%) | 2 (33.3%) |
| Eyesight abnormality | n (%) | 2 (40%) | 1 (16.7%) | 2 (28.6%) | 6 (31.6%) | 0 (0%) |
| AUDIT score | mean (±SD) | 22.5 (±8.1) | 21.7 (±1.5) | 16.3 (±5.9) | 20.1 (±6.8) n=13 | 11.7 (±3.2) n=3 |
| PAE length weeks | mean (±SD) | 24.2 (±10.8) | 16.2 (±12.5) | 10.1 (±7.8) | 15.4 (±11.4) | 12.2 (±6.4) |
| Early only exposure | n (%) | 0 (0%) | 3 (50%) | 4 (57.1%) | 8 (42.1%) | 2 (33.3%) |

Finally, we further examined the timing of maternal alcohol consumption in relation to diagnoses. We defined the duration of early exposure up to the GW 7 at maximum, as in our previous study (Auvinen et al., 2022). In the early PAE group, heavy alcohol consumption was more prevalent compared to longer PAE (Supplementary Figure S2). Regarding the diagnoses, none of the children in the early PAE subgroup was diagnosed with FAS, three had PFAS, and four met the criteria of ARND. Neither the children in the early PAE subgroup (*n*=11) nor all other children in the longer PAE subgroup (*n*=17) differed significantly in anthropometric measurements from the control children at birth, when GA and sex were considered (SDs) (Supplementary Table S9a). However, in the longer PAE group, significantly more six-year-olds had been underweight, overweight, or had smaller HCs compared to the controls (≤ −1.25 SD) (OR 4.5, 95% CI 1.1-18.6, *P*<0.05; OR 5.8, 95% CI 1.3-25.3, *P*<0.05; OR 10.2, 95% CI 2.2-48.6, *P*<0.01), respectively).

Upon further analysis of the timing of PAE and associated dysmorphology, similar rates of dysmorphic features were observed in both the early and longer exposure groups (Supplementary Table S9b). In terms of cognitive performance, half of the PAE children (4/8, 50%) with borderline or lower FSIQ scores (FSIQ < 85) were members of the early PAE group (Supplementary Table S9c).

## DISCUSSION

### Dysmorphological and neuropsychological features associated with PAE

As expected, children with PAE had significantly more dysmorphic features and both neurobehavioral and somatic problems than controls. At the age of six, 19/25 (76%) of those with PAE fulfilled FASD criteria, and five out of 25 met the criteria of FAS. Dysmorphological assessment showed that PAE causes a variety of dysmorphological changes in addition to the well-recognized facial features of FAS. A negative correlation between dysmorphology scores and both HC and weight (SD) at birth and at the age of six indicates that the greater dysmorphology severity is linked to lower birth weight and smaller HC, both indicative of fetal developmental impairment resulting from PAE.

Unexpectedly, PAE was associated not only with underweight but also with an increased risk of overweight among six-year-olds, particularly among boys. Previously, it has been shown that adolescent girls with FASD have a heightened risk of overweight (Fuglestad et al., 2014), and that abnormal eating behaviors and impaired satiety mechanisms are common among children with FASD (Amos-Kroohs et al., 2016), potentially contributing to the development of overweight. However, given the limited sample size, these findings should be validated in future studies.

Children with PAE showed lower average performance on cognitive tests compared to the control group, and a total of seven out of 25 met the criteria of ARND, having only the neurobehavioral phenotype of FASD. ADHD-related symptoms represented the most common form of neurobehavioral impairment. This is consistent with previous studies indicating that ADHD is the most prevalent symptom associated with FASD (Burd, 2016; Clark et al., 2024). The earliest age at which ADHD can be diagnosed with reasonable reliability is approximately six years, and most individuals with ADHD related to PAE receive their diagnosis at a later stage (Clark et al., 2024). This highlights the importance of identifying both FASD and other PAE-related difficulties that may not yet meet the formal FASD criteria at that age.

### Diagnostics of FASD

Owing to the heavy exposure of the children in the PAE group, the high prevalence of FASD is not surprising. It is likely that some children who have not yet received a formal diagnosis will exhibit pronounced symptoms of FASD, as learning disabilities, executive function deficits, and behavioral challenges become more evident with advancing age and the increasing demands of school and societal expectations. The age of six has both advantages and disadvantages in assessing the effects of PAE. Many in-school FASD prevalence studies assess children in the first grade (approximately ages 6–7). This can be considered an early diagnosis that enables planning support for the beginning of school. Children are typically able to cooperate during testing at this age, and attention problems are beginning to emerge. However, the nurturing environment characteristic of this developmental stage can offset some of the cognitive difficulties associated with PAE, as abstract reasoning and independence in many adaptive skills are not yet required or evaluated in neuropsychological assessments.

Our results confirm the association of PAE with cognitive problems and ADHD symptoms, which have been included in the IOM 2016 diagnostic criteria of FASD (Hoyme et al., 2016). However, several PAE-associated features, which are not included in that diagnostic criteria, such as reduced postural stability and lower performance in static balance tasks (Lucas et al., 2014), were also observed in the current study. PAE increases risks for many dysmorphologies as well as ear infections, very selective eating, and problems in eyesight, self-help skills, and social functioning, which neither included in the diagnostic criteria (Attell et al., 2025; Popova et al., 2016). Even though adaptive functioning is not included in used IOM 2016 diagnostic criteria for FASD, it has been shown to be more affected by PAE than IQ scores in neuropsychological testing (Fagerlund et al., 2012; Streissguth et al., 2004). However, both adaptive and social functioning problems have been included in the DSM-5 ND-PAE (Neurobehavioral Disorder associated with PAE), which may capture symptoms of ARND better than the current IOM 2016 criteria.

Several different diagnostic guidelines for FASD are in use (Myers et al., 2025; Hemingway et al., 2019), and current diagnostic guidelines predominantly emphasize the characteristic facial features of FAS, due to their relative specificity to the disorder. However, it has been shown that a single ethanol exposure during mouse gastrulation – which corresponds to the fifth week of human gestation – can cause craniofacial abnormalities similar to those seen in humans with FAS (Lipinski et al., 2012). Considering this as well as the high prevalence of ARND, concentrating only on certain dysmorphological traits may fail to capture important outcomes of exposure. Since PAE before and after gastrulation leads to different types of facial anomalies as well as changes in neurodevelopment and brain morphology (Lipinski et al., 2012; Muggli et al., 2024), the typical FAS facial features do not reflect the full extent of dysmorphology, or other developmental disorders caused by exposure. As a result, the choice of diagnostic criteria can affect both the proportion and the phenotype of PAE children identified with FASD, highlighting the importance of comprehensively reporting the phenotypic characteristics observed in the PAE population relative to unexposed controls.

### Effects of early exposure on phenotype

When the timing of maternal alcohol consumption in relation to the diagnoses was examined, none of the six-year-old children exposed to alcohol up to GW 7 were diagnosed with FAS; instead, they were diagnosed with PFAS or ARND. One explanatory factor is the restricted growth that is required for an FAS diagnosis, and there were no children meeting the criteria for growth impairment in the early exposure group. Interestingly, they had similar rates of other dysmorphic features compared to the longer exposure group, and four out of eight children in the PAE group with low full-scale IQ scores (FSIQ < 85) were in the early exposure subgroup. This emphasises the vulnerability of early pregnancy to alcohol exposure, particularly the sensitivity of the nervous system, and is consistent with earlier studies (Legault et al., 2021; Wallén et al., 2025). This is also consistent with the guidelines recommending the use of contraception when consuming alcohol and advising individuals to stop drinking alcohol when planning a pregnancy (Carson et al., 2010). Recently, it has been shown that as a membrane-permeable molecule, ethanol can influence embryonic and placental development even prior to implantation (Legault et al., 2021). It is also worth noting that the mean maternal AUDIT score in the early PAE group was 21.3, indicating heavy exposure.

### Limitations of the study

We recognize the limitations of this study. The sample size was relatively small, which reduced the statistical power and sex-specific analyses were not feasible to perform. Furthermore, given the previously observed sex-specific effects of PAE, it is noteworthy that the PAE group included a higher proportion of females (17 of 28; 61%) compared with the control group (25 of 52; 48%), which may have influenced some results. In addition, self-reported information about the amount of alcohol consumed and the duration of use may be inaccurate, as many mothers underreport their consumption due to stigma or fear of social repercussions (Gomez-Roig et al., 2018; Lange et al., 2014). However, considering that the length of PAE and AUDIT scores correlated with dysmorphology and SRS scores, respectively, and that there was an absence of growth impairment in the early PAE group, the results support the reliability of the self-reported information.

The study may have participant selection bias, as control parents with some concern about their child’s development are more likely to participate than those without concerns. On the other hand, parents with ongoing serious substance abuse and stressful everyday lives are difficult to reach and engage. Additionally, many children who already had support actions in place were ineligible to participate. However, when the representativeness of the tested children relative to the cohort was assessed, there were no significant differences between the untested and tested groups. Considering the higher number of small HCs (HC ≤ −1.25 SD) and other malformations observed in the untested PAE children, this group is unlikely to have been exposed to a lower amount of alcohol or to consist of less affected children. Based on this, we can conclude that the group of children tested is well representative of the epiFASD cohort and the majority of PAE children in the whole cohort would have an FASD diagnosis.

Six years of age may not be optimal for certain evaluations. The tests were conducted before school entry, and many learning difficulties as well as attention or executive function deficits often do not become apparent until formal schooling begins and demands for independent functioning increase. Also, at the age of six, differences in parental nurture and other environmental differences may still have a strong impact on children’s cognitive test results. Adoption and twin studies have shown that during early childhood, environmental factors have significantly more impact on IQ than in later life, when congenital factors outweigh them (Haworth et al., 2010; Bouchard, 2013). It is also important to acknowledge the influence of genetic background, especially due to reported attention difficulties and ASD among family members. Due to the limited sample size, these aspects could not be examined in the current study, but they represent important issues to be addressed in future research.

## CONCLUSIONS

This study confirms previously established associations between PAE and dysmorphological features, cognitive impairments, and ADHD symptoms in children. We also observed additional phenotypic features that are not included in the current FASD diagnostic criteria, such as increased risks for somatic health conditions as well as difficulties in adaptive and social functioning. These results highlight the complexity of the FASD phenotype and point to the importance of continued efforts to refine diagnostic methods to adequately identify the full range of children affected by alcohol exposure. Finally, our results emphasize the vulnerability of early embryonic development to alcohol, as even exposure limited to the first seven weeks of pregnancy was associated with dysmorphological and neurocognitive impairments at the age of six.

## Data Availability

All data produced in the present work are contained in the manuscript.

## ACKNOWLEDGEMENTS

We are grateful to all the families who took part in this study. We would also like to acknowledge research nurses Jaana Palukka and Teija Karkkulainen for their valuable contribution to this work.

